# Protection from Omicron infection in residents of nursing and retirement homes in Ontario, Canada

**DOI:** 10.1101/2022.06.28.22277016

**Authors:** Jessica A. Breznik, Ahmad Rahim, Tara Kajaks, Megan Hagerman, Lucas Bilaver, Karen Colwill, Roaya M. Dayam, Anne-Claude Gingras, Chris P. Verschoor, Janet E. McElhaney, Jonathan L. Bramson, Dawn M. E. Bowdish, Andrew P. Costa

## Abstract

**Objectives:** To identify factors that contribute to protection from infection with the Omicron variant of SARS-CoV-2 in older adults in nursing and retirement homes.

**Design:** Longitudinal cohort study with retrospective analysis of infection risk.

**Setting and Participants:** 997 residents of nursing and retirement homes from Ontario, Canada, in the COVID-in-LTC study.

**Methods:** Residents with three mRNA dose vaccinations were included in the study. SARS-CoV-2 infection was determined by positive nasopharyngeal PCR test and/or circulating anti-nucleocapsid IgG antibodies. Cumulative probability of Omicron infection after recent COVID-19 was assessed by log-rank test of Kaplan-Meier curves. Cox regression was used to assess risk of Omicron infection by age, sex, mRNA vaccine combination, whether individuals received a fourth dose, as well as recent COVID-19.

**Results:** 171 residents (17.2%) had a presumed Omicron variant SARS-CoV-2 infection between December 15, 2021 (local start of the first Omicron wave) and May 3, 2022. Risk of Omicron infection was not different by age [hazard ratio (95% confidence interval): 1.01 (0.99-1.02)], or in women compared to men [0.97 (0.70-1.34)], but infection risk decreased 47% with three vaccine doses of mRNA-1273 (Moderna) compared to BNT162b2 (Pfizer) [0.53 (0.31-0.90)], 81% with any fourth mRNA vaccine dose [0.19 (0.12-0.30)], and 48% with SARS-CoV-2 infection in the 3 months prior to beginning of the Omicron wave [0.52, (0.27-0.99)].

**Conclusions and Implications:** Vaccine type (i.e., mRNA-1273/Spikevax vs BNT162b2/Cominarty), any fourth vaccine dose, and hybrid immunity from recent COVID-19, were protective against infection with the Omicron variant. These data emphasize the importance of vaccine type, and number of vaccine doses, in maintenance of protective immunity and reduction of risk of Omicron variant breakthrough infection. These findings promote continued public health efforts to support vaccination programs and monitor vaccine immunogenicity in older adults.

**Brief summary:** Risk of infection with the SARS-CoV-2 Omicron variant in older adults in early 2022 was reduced with triple mRNA-1273 vaccination, any fourth dose vaccine, and within three months of prior COVID-19.

## Introduction

The highly transmissible and immune evasive Omicron (B.1.1.529) SARS-CoV-2 variant of concern has caused an unprecedented and precipitous global rise in COVID-19 cases^1,2^. In the province of Ontario, Canada, the Omicron variants BA.1 and BA.2 dominated the fifth and sixth waves of infections from December 2021, extending into Spring/Summer 2022^3^. Waning protective immunity from vaccination, in combination with reduced neutralization capacity of vaccine-elicited antibodies against the Omicron variant^4,5^, may have contributed to widespread outbreaks and a corresponding rise in hospitalization and deaths, particularly in older adults^6^. However, compared to earlier SARS-CoV-2 variants, Omicron generally causes less severe illness in community-dwelling adults^7,8^, and even residents of nursing homes^9^. This is likely due to high rates of vaccination^10^ and/or hybrid immunity due to infections with earlier variants^11,12^.

Prior to the emergence of the Omicron variant, most older adults in nursing and retirement homes in Ontario had primary series (i.e. two doses) and third dose vaccinations with mRNA vaccines^3^, with ∼86% of eligible nursing home residents having received a third dose as of December 8, 2021^13^. We previously found waning antibody levels and neutralization capacity within several months after second and third mRNA vaccines in nursing and retirement home residents^14,15^. As third doses were recommended for these populations in Ontario as of August 17, 2021^16^, many individuals were several months from their most recent SARS-CoV-2 vaccine when Omicron infections began spreading in the community. Given this, concerns about the longevity of protective immunity after third vaccine doses in older adults in congregate living facilities prompted recommendations for fourth doses^13^, with rollout in early 2022. However, COVID-19 outbreaks and enactment of infection prevention and control measures may have prevented timely vaccination of many individuals, thereby increasing infection risk. In addition, while residents of nursing and retirement homes in Ontario received mRNA vaccines almost exclusively, some received mixes of vaccine brands. Based on what we and others have reported, there is enhanced vaccine immunogenicity of the mRNA-1273 (Moderna/Spikevax) vaccine compared to BNT162b2 (Pfizer/Cominarty)^14,17,18^, and many, but not all, residents received mRNA-1273 for their third and fourth doses.

We hypothesized that differences in the number of mRNA vaccine doses and vaccine type combinations could be contributing factors to Omicron infection susceptibility in older adults. As well, humoral vaccine responses are often increased in people who have recovered from COVID-19^11,12,19^, so we predicted that individuals with a recent SARS-CoV-2 infection may have protective hybrid immunity against the Omicron variant. In a cohort of residents of nursing and retirement homes, we examined factors including age, sex, vaccine combination, prior infection, and whether individuals received fourth vaccine doses, to determine which contributed to protection from infection with the Omicron variant of SARS-CoV-2.

## Methods

Participants in the COVID in Long-Term Care Study (https://covidinltc.ca/) were recruited from 17 nursing homes and 8 retirement homes in Ontario, Canada, between March and November 2021. All protocols were approved by the Hamilton Integrated Research Ethics Board and other site-specific research ethics boards, and informed consent was obtained. Analyses in this article were performed with data from 997 participants that were alive and had received three mRNA vaccine doses as of December 15, 2021. Participants received 2 doses of Moderna Spikevax 100 µg (mRNA-1273) or Pfizer Cominarty 30 µg (BNT162b2) as per recommended schedules, and a third mRNA vaccine dose in Fall 2021 at least 6 months from the last dose, as per Province of Ontario guidelines^20^. Fourth doses vaccinations were available as of early 2022^13^. Due to the demand for and limits on PCR testing, lack of routine screening for SARS-CoV-2 amongst residents, and observations that Omicron infections may have a higher rate of asymptomatic carriage^21^, COVID-19 infections between December 15, 2021 (the start of the SARS-CoV-2 Omicron variant wave as defined by Public Health Ontario)^22^ and May 3, 2022, were determined by record of confirmed PCR test positivity and/or dried blood spot (DBS) seropositivity for anti-nucleocapsid IgG antibodies by validated ELISA^23^. DBS were collected from finger pokes or venous blood at 2-6 weeks, 2-4 months, and/or 5-7 months after second and third dose vaccinations, and 2-4 months after fourth dose vaccinations, if applicable. Analyses were performed with a relative ratio (raw counts normalized to a point on a synthetic antibody reference curve included in each assay plate) seropositivity threshold of 0.350 (specificity of 89.2% and sensitivity of 96.9% using samples obtained pre-COVID-19 or from unvaccinated convalescent individuals, defined in^23^), and supplementary analyses were performed with threshold seropositivity of 0.642 (specificity of 99.0% and sensitivity of 92.0%). The first DBS to pass the threshold of seropositivity for anti-nucleocapsid IgG antibodies was identified as a primary infection. Positive DBS after this point were considered part of the same infection, until a decline in assay measurement was observed, indicative of a decrease or waning of antibodies. After this decline, a subsequent increase in signal that was above the seropositivity threshold, and at least 20% greater than the previous DBS, was assumed to be caused by a subsequent new infection. Positive DBS and/or PCR that were within 30 days of each other were considered part of the same infection, and the earlier associated date was used as the estimated date of infection. Any reported positive PCR tests that were not within 30 days of a seropositive DBS or other positive PCR test were considered a separate infection. Province-wide genome sequencing identified that the Omicron variant accounted for over half of infections in Ontario as of December 13, 2021, and over 90% of infections as of December 23, 2021^24^. Infections in our cohort from December 15, 2021 onward were therefore presumed to be caused by the Omicron variant as genetic strain verification testing was unavailable.

Differences in baseline characteristics between individuals that were infected with Omicron and not infected with Omicron were assessed by chi-square test of independence (proportions), Student’s t-test (mean), and Mann-Whitney U test (median). Kaplan-Meier curves were generated to show the cumulative incidence of Omicron infection over time by baseline previous infection status (i.e., no infection within three months prior to December 15, 2021 or infection within three months), determined as described above by serial DBS and/or PCR testing. The log-rank test was used to determine statistical difference between the curves. Right censoring occurred when participants died, declined to continue in the study, or did not develop the outcome at the end of the follow-up period. Cox regression was used to estimate the hazard ratios of Omicron infection by baseline previous infection status, with the baseline hazard on December 15, 2021. Variables (age, sex, mRNA vaccine combination received, and previous infection status) reflect time to baseline. To estimate the change in risk of Omicron infection due to receiving a fourth mRNA vaccine during the follow-up period, we included a binary time-dependent covariate (i.e., received a fourth vaccine or did not receive a fourth vaccine) in the regression analysis. Regression results are presented as hazard ratios with 95% confidence intervals based on robust standard errors, accounting for the clustering of participants by facility. A P-value of < 0.05 was considered statistically significant. All statistical analyses were performed using SAS v9.4 (SAS Institute Inc) and Kaplan-Meier curves were plotted with R v4.2.0 (R Core Team).

## Results

In our cohort of 997 residents of nursing and retirement homes, 17.2% (n=171) had a breakthrough infection between December 15, 2021 and May 3, 2022 (Table 1). Baseline characteristics of age and sex were not different between individuals with infection (median 87.0 years, IQR 80.0-92.0; 66.1% female) and without infection (median 87.0 years, IQR 80.0-91.0; 66.7% female). However, there was a decreased frequency of infections in individuals with three mRNA-1273 vaccines, or any other triple mRNA vaccine combination, compared to three BNT162b2 vaccines (P < 0.001; infection incidence of 27.5% (47/171) 3x mRNA-1273, 4.68% (8/171) 3x mRNA combination, 67.8% (116/171) 3x BNT162b2). Individuals who had a SARS-CoV-2 infection in the three months prior to the emergence of the Omicron variant had a lower incidence of Omicron infection (P < 0.001; 6.43% (11/171) infection, 93.6% (160/171) no infection). Further investigation identified that the cumulative probability of being infected with the Omicron variant was lower in individuals with recent SARS-CoV-2 infection (P < 0.001) (Figure 1).

**Table 1.** Characteristics of study cohort.

|  | *Total* | *Omicron Infection?* |  | *P* |
| --- | --- | --- | --- | --- |
|  | *(N = 997)* | *Yes*<br>*(N = 171)* | *No*<br>*(N = 826)* |  |
| **Age** |  |  |  |  |
| Mean (SD) | 84.6 (9.94) | 84.9 (9.17) | 84.5 (10.1) | 0.59 |
| Median (IQR) | 87.0 (80.0 – 91.0) | 87.0 (80.0 – 92.0) | 87.0 (80.0 – 91.0) | 0.70 |
| **Sex** |  |  |  |  |
| Female | 664 (66.6%) | 113 (66.1%) | 551 (66.7%) | 0.87 |
| Male | 333 (33.4%) | 58 (33.9%) | 275 (33.3%) |  |
| **Vaccine combination** |  |  |  |  |
| mRNA-1273 x3 | 420 (42.1%) | 47 (27.5%) | 373 (45.2%) | <0.001 |
| BNT162b2 x3 | 478 (47.9%) | 116 (67.8%) | 362 (43.8%) |  |
| mRNA combination x3 | 99 (9.93%) | 8 (4.68%) | 91 (11.0%) |  |
| **COVID-19 in past 3 months** |  |  |  |  |
| No | 853 (85.6%) | 160 (93.6%) | 693 (83.9%) | 0.001 |
| Yes | 144 (14.4%) | 11 (6.43%) | 133 (16.1%) |  |

**Figure 1.**
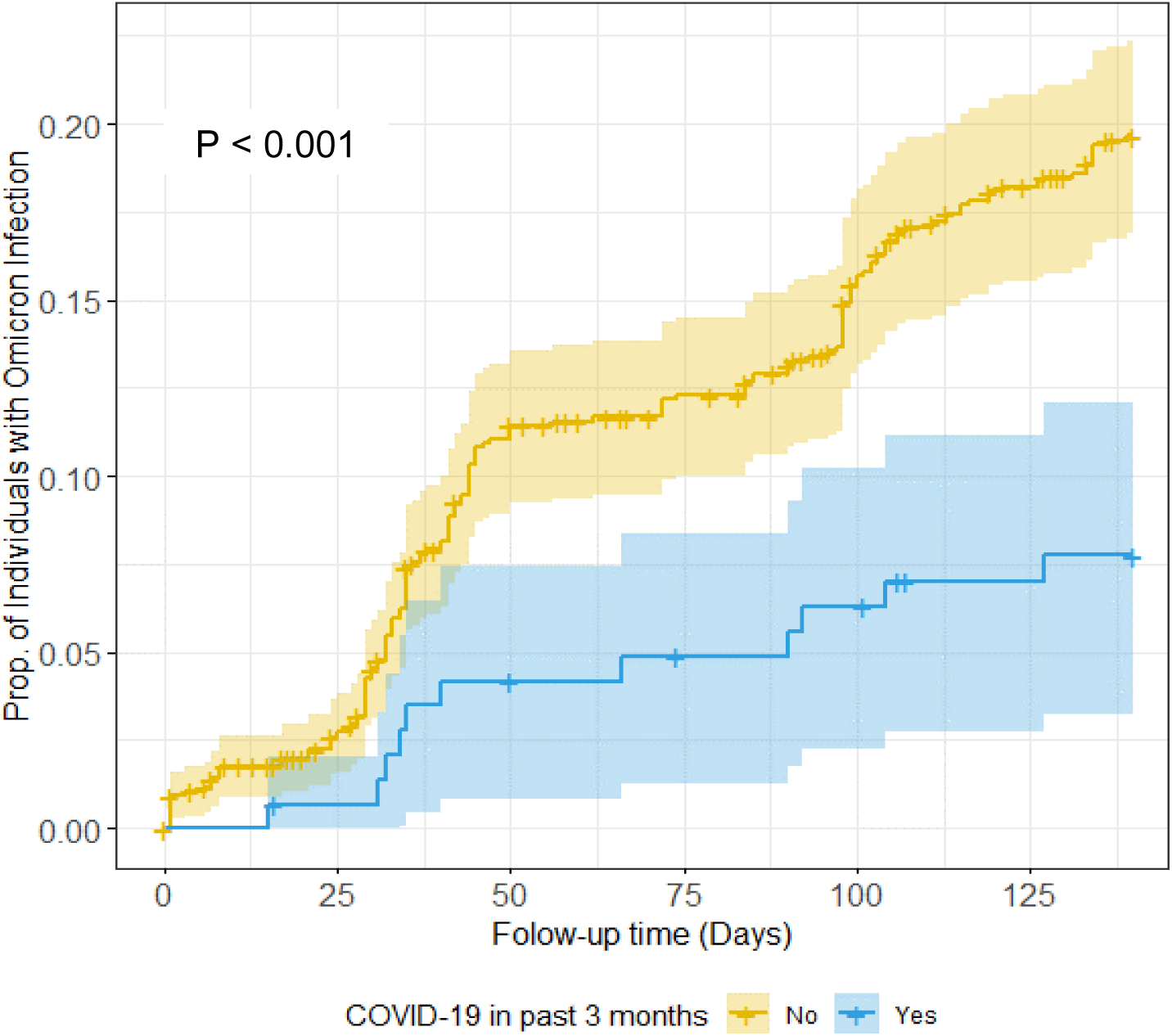
Kaplan-Meier curves of incidence of Omicron infection in nursing and retirement homes by recent SARS-CoV-2 infection.

We used multivariate regression to estimate risk of Omicron infection (Figure 2; Supplementary Table 1). We found that age and sex had little effect on risk of infection, but vaccine type, number of vaccines, and prior infection, modulated risk. Risk of Omicron infection was significantly decreased in individuals who received three mRNA-1273 vaccines compared to three BNT162b2 vaccines [hazard ratio (95% confidence interval): HR 0.53 (0.31-0.90)]. Individuals with any fourth vaccine dose likewise had decreased risk of Omicron [HR 0.19 (0.12-0.30)]. Omicron infection risk was also decreased in individuals who had COVID-19 in the 3 months prior to December 15, 2021 [HR 0.52, (0.27-0.99)]. Therefore, in residents of nursing and retirement homes, vaccine combination as well as recent vaccination or COVID-19, influenced risk of Omicron infection.

**Figure 2.**
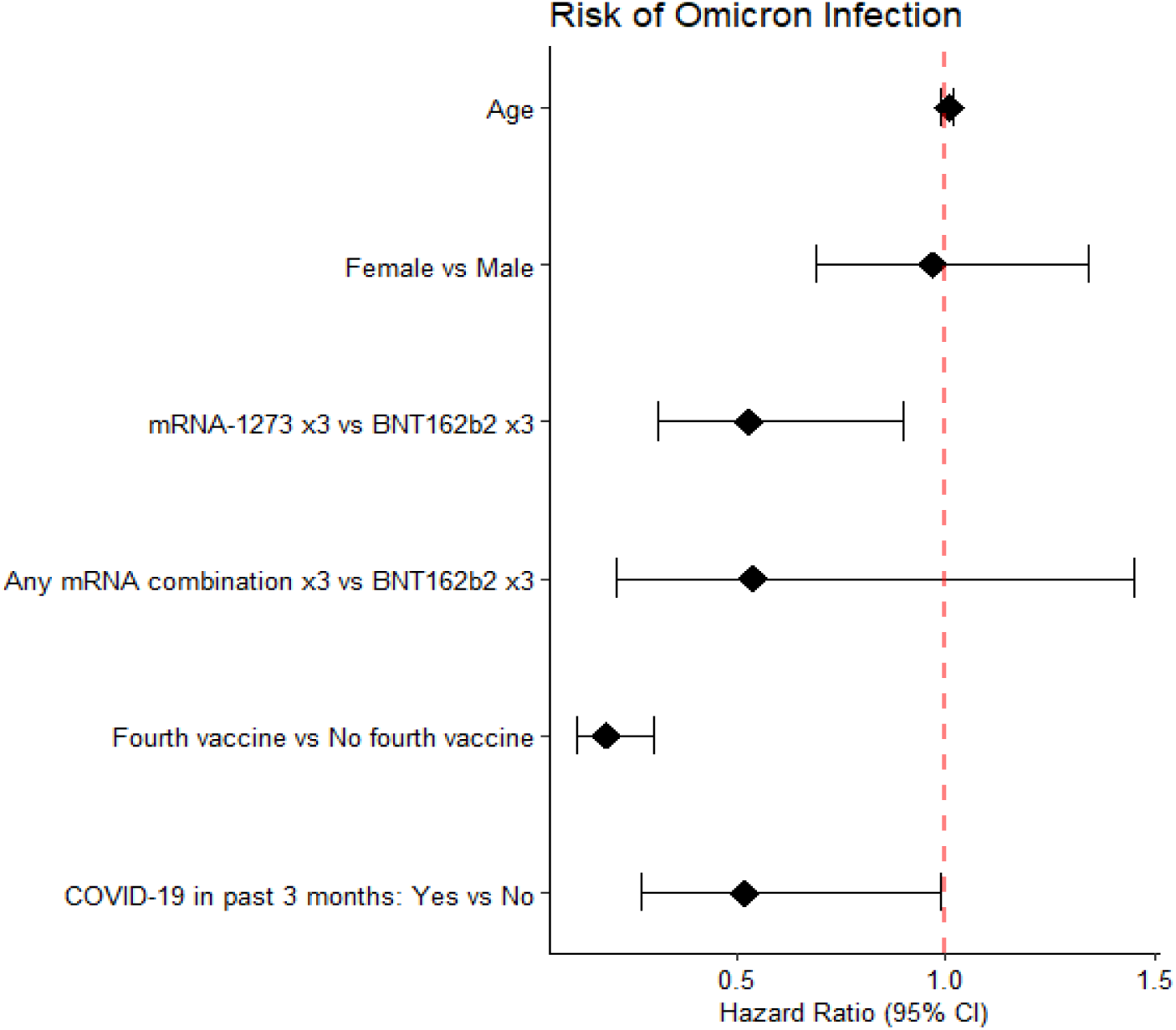
Cox regression of factors contributing to risk of Omicron infection in nursing and retirement homes.

## Discussion

In this investigation of SARS-CoV-2 infection in nursing and retirement home residents, we identified that vaccine type, a fourth dose, and hybrid immunity from recent COVID-19, influenced risk of infection with the Omicron variant. Congruent with our prior observations of higher humoral immunogenicity of mRNA-1273 vaccines compared to BNT162b2 vaccines after two vaccine doses^14^, we found that three doses of mRNA-1273 conferred a 52.4% lower risk of Omicron infection. This could be due to a higher mRNA dose, and/or differences in humoral and T cell responses that have been recently described in community-dwelling adults^14,17,18,25^.

Importantly, we found that any fourth mRNA vaccine was protective against Omicron infection, with 81% reduced risk. This observation suggests that in older adults, vaccine-elicited immunity from a third vaccine wanes within several months. This is consistent with what is known about the immune-evasive properties of the Omicron variant and the accelerated waning of humoral responses that occurs in older adults. Vaccination at the onset of new waves of infection will be essential to ensure sufficiently high antibody levels to protect from infection^4,5^. These observations align with studies in older adults and the general population that report individuals with a recent third vaccination were at lower risk of hospital admission due to Omicron^7,8^. Consequently, maintaining optimal protection through vaccination, irrespective of infection history, is important for longevity of immune protection. Our findings are especially important to consider in context of similar populations like community-dwelling older adults, who may not have the same incidence of recent infections, nor comparable levels of vaccine coverage.

The combination of SARS-CoV-2 vaccination and natural infection, or hybrid immunity, influences risk of COVID-19 incidence and severity^26,27^. A strength of our study is that we did not rely on symptomatic PCR determination of SARS-CoV-2 infection, but also incorporated serological determination of recent infection, which made it possible to identify presumptively asymptomatic infections when a positive PCR test was not reported. Our analyses show that hybrid immunity conferred a 48% lower risk of Omicron infection in older adults in congregate living facilities. It seems probable that a recent SARS-CoV-2 infection in older adults would have provided a ‘boost’ to immunity, even in the absence of a fourth vaccine dose. It will be of interest to further examine the longevity of this apparent hybrid protection, as it has been reported that anti-nucleocapsid IgG levels wane to undetectable levels in half of older adults within 8 months^28^. Furthermore, as infections were dominated by the Delta variant in Ontario in the months preceding emergence of the Omicron variant, it is unknown whether infection with Omicron protects from subsequent Omicron infection. Consecutive Omicron infections have been recently reported^29,30^, perhaps due to gradual changes in circulating subvariants of Omicron, or due to low protective immunity after vaccination, the latter of which may be of particular concern in older adults.

This study has several limitations. As we used both symptomatic PCR testing and DBS measurements of serum anti-nucleocapsid IgG to determine SARS-CoV-2 infection incidence, most dates of infection were estimated from dates of DBS collection. Though the use of DBS allowed detection of presumptive asymptomatic or mild infections in the absence of a positive PCR test, antibody production may in particular be lower after asymptomatic infection. For this reason, we chose a low threshold for seropositivity in our assessments, but similar parameters were identified to influence infection risk when analyses were repeated with a higher antibody seropositivity threshold (Supplementary Figure 1, Supplementary Tables 2 and 3). Furthermore, risk analyses were not stratified according to severity of infection or infection outcomes, and we did not have access to genomic sequencing data in symptomatic PCR-positive cases, which prevented confirmation of infection with the Omicron variant.

## Conclusions and Implications

We found that risk of Omicron infection was significantly lower in residents with three mRNA doses of mRNA-1273 compared to BNT162b2, a fourth dose of any mRNA vaccine, or hybrid immunity from COVID-19 within three months of the emergence of the Omicron variant. Our observations add to accumulating evidence of the relevance of recent vaccination and infection in susceptibility to the Omicron variant, and more broadly SARS-CoV-2. These data accentuate the importance of maintenance of protective immunity in older adults to prevent SARS-CoV-2 infection. Moreover, our data emphasize that continued epidemiological and serological surveillance by public health agencies is essential to assess long-term immunogenicity of vaccines, longevity of hybrid immunity, and the need for vaccination and other infectious disease control measures in vulnerable older adult populations.

## Data Availability

All data produced in the present study are available upon reasonable request to the authors.

## Author Contributions

Drs Bowdish and Costa had full access to all of the data in the study and take responsibility for the integrity of the data and the accuracy of the data analysis.

*Study concept and design:* Bowdish, Costa.

*Acquisition, analysis, or interpretation of data:* All authors.

*Drafting of the manuscript*: Breznik.

*Critical revision of the manuscript for important intellectual content:* All authors.

## Additional Information

Data in this study were collected by the COVID-in-LTC Study Group. Members of the COVID-in-LTC Study Group include Eric D. Brown, PhD; Kevin Brown, PhD; David C. Bulir, MD, PhD; Judah A. Denburg, MD; George A. Heckman, MD, MSc; Michael P. Hillmer, PhD; John P. Hirdes, PhD; Aaron Jones, PhD; Mark Loeb, MD, MSc; Matthew Miller, PhD; Ishac Nazy, PhD; Nathan M. Stall, MD; Parminder Raina, PhD; Marek Smieja, MD, PhD; Kevin J. Stinson, PhD; Ahmad Von Schlegell; Arthur Sweetman, PhD; Gerry Wright, PhD.

## Additional Contributions

We would like to thank our participants, their families, as well as partner homes and staff, for their support of this study. We acknowledge administrative and technical assistance from members of the COVID-in-LTC Study Group, who were compensated for their contributions by a grant funded by the Canadian COVID-19 Immunity Task Force at McMaster University. Nucleocapsid ELISAs were performed at the Network Biology Collaborative Centre at the Lunenfeld-Tanenbaum Research Institute, a facility supported by Canada Foundation for Innovation funding, by the Government of Ontario and by Genome Canada and Ontario Genomics (OGI-139). We thank Geneviève Mailhot, MSc, Melanie Delgado-Brand, Tulunay Tursun and Freda Qi for assistance with the ELISAs.

