## Supplemental Figure 1 and Tables 1-3 for "Protection from Omicron infection in residents of nursing and retirement homes in Ontario, Canada"

**Supplementary Figure 1. Alternate antibody threshold Kaplan-Meier curves and Cox regression analyses of risk of Omicron infection in nursing and retirement homes.**

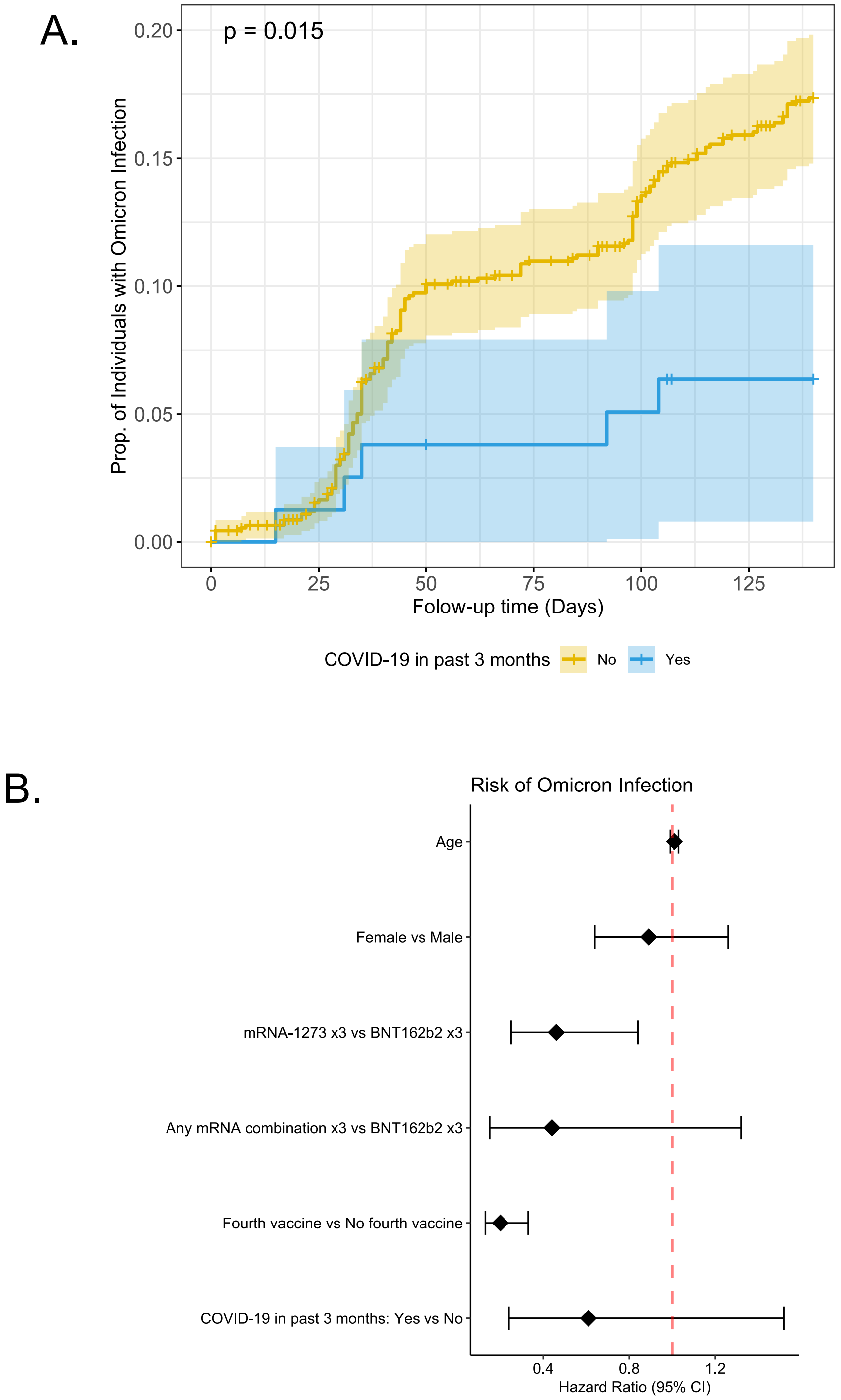


(A) Kaplan-Meier curve of incidence of Omicron infection in nursing and retirement homes by recent SARS-CoV-2 infection. (B) Cox regression of factors contributing to risk of Omicron infection in nursing and retirement homes. Anti-nucleocapsid seropositivity threshold was set at 0.642.

**Supplementary Table 1. Analysis of maximum likelihood estimates of risk of Omicron infection in nursing and retirement homes.**

| Parameter | Estimate | Standard Error | Chi-Square | Pr > ChiSq | Hazard Ratio | 95% Hazard Ratio Confidence Limits | |
|---|---|---|---|---|---|---|---|
| **Age** | 0.0057 | 0.0088 | 0.43 | 0.51 | 1.01 | 0.99 | 1.02 |
| **Sex** (Female vs Male) | -0.035 | 0.17 | 0.044 | 0.83 | 0.97 | 0.70 | 1.34 |
| **Vaccine Type** (mRNA-1273 x3 vs BNT162b2 x3) | -0.63 | 0.27 | 5.48 | 0.019 | 0.53 | 0.31 | 0.90 |
| **Vaccine Type** (mRNA x3 vs BNT162b2 x3) | -0.61 | 0.50 | 1.49 | 0.22 | 0.54 | 0.21 | 1.45 |
| **Fourth mRNA Vaccine** (Yes vs No) | -1.65 | 0.23 | 49.82 | <0.001 | 0.19 | 0.12 | 0.30 |
| **COVID-19 in past 3 months** (Yes vs No) | -0.66 | 0.33 | 3.96 | 0.047 | 0.52 | 0.27 | 0.99 |

**Supplementary Table 2. Alternate antibody threshold study cohort characteristics.**

| *Antibody Threshold 0.642* | | | | |
|---|---|---|---|---|
| | ***Total*** | ***Omicron Infection?*** | | ***P*** |
| | | ***Yes*** | ***No*** | |
| | ***(N = 987)*** | ***(N = 152)*** | ***(N = 835)*** | |
| **Age** | | | | |
| Mean (SD) | 84.6 (9.94) | 85.2 (9.00) | 84.4 (10.1) | 0.41 |
| Median (IQR) | 87.0 (80.0 – 91.0) | 87.0 (80.0 – 92.0) | 87.0 (80.0 – 91.0) | 0.54 |
| **Sex** | | | | |
| Female | 664 (66.6%) | 102 (65.0%) | 562 (66.9%) | 0.64 |
| Male | 333 (33.4%) | 55 (35.0%) | 278 (33.1%) | |
| **Vaccine combination** | | | | |
| mRNA-1273 x3 | 420 (42.1%) | 36 (22.9%) | 384 (45.7%) | <0.001 |
| BNT162b2 x3 | 478 (47.9%) | 114 (72.6%) | 364 (43.3%) | |
| mRNA combination x3 | 99 (9.93%) | 7 (4.46%) | 92 (11.0%) | |
| **COVID-19 in past 3 months** | | | | |
| No | 918 (92.1%) | 152 (96.8%) | 766 (91.2%) | 0.017 |
| Yes | 79 (7.92%) | 5 (3.18%) | 74 (8.81%) | |

**Supplementary Table 3. Alternate antibody threshold analysis of maximum likelihood estimates of risk of Omicron infection in nursing and retirement homes.**

| Antibody Threshold 0.642 | | | | | | | |
|---|---|---|---|---|---|---|---|
| **Parameter** | **Estimate** | **Standard Error** | **Chi-Square** | **Pr > ChiSq** | **Hazard Ratio** | **95% Hazard Ratio Confidence Limits** | |
| **Age** | 0.012 | 0.0094 | 1.54 | 0.22 | 1.01 | 0.99 | 1.03 |
| **Sex** (Female vs Male) | -0.11 | 0.17 | 0.39 | 0.54 | 0.90 | 0.64 | 1.26 |
| **Vaccine Type** (mRNA-1273 x3 vs BNT162b2 x3) | -0.78 | 0.31 | 6.31 | 0.012 | 0.46 | 0.25 | 0.84 |
| **Vaccine Type** (mRNA x3 vs BNT162b2 x3) | -0.82 | 0.56 | 2.16 | 0.14 | 0.44 | 0.15 | 1.32 |
| **Fourth mRNA Vaccine** (Yes vs No) | -1.60 | 0.25 | 42.89 | <0.001 | 0.20 | 0.13 | 0.33 |
| **COVID-19 in past 3 months** (Yes vs No) | -0.50 | 0.47 | 1.14 | 0.29 | 0.61 | 0.24 | 1.52 |
